# Clinical features, first aid practices, and hospital management of snakebites in Northern Uganda: a multi-facility cross-sectional study

**DOI:** 10.64898/2026.07.07.26357437

**Authors:** Solomon T. Wafula, Akello Trophy, Shakiim Mubiru, Richard K Mugambe, John Bosco Isunju, John Bosco Ddamulira, David Musoke

## Abstract

Snakebite envenoming is a neglected tropical disease with a disproportionate burden in sub-Saharan Africa, where weak surveillance, limited antivenom availability, and reliance on traditional remedies compound morbidity and mortality. Evidence on how snakebite cases are clinically managed in routine practice in Uganda remains scarce. We conducted a facility-based cross-sectional study with retrospective abstraction of records from four purposively selected health facilities in Gulu and Arua districts of Northern Uganda. Snakebite case records from outpatient, inpatient, paediatric, and intensive-care service points covering the period 2017–2021 were reviewed using a standardised abstraction tool. Descriptive statistics were computed in Stata version 14, with frequencies and proportions for categorical variables and medians with interquartile ranges (IQR) for continuous variables. A total of 227 snakebite case records were analysed, of which 58.6% were female, a median age of 21 years (IQR 13–38), and 31.3% were students. Most bites involved the lower limb (85.9%) and occurred in the second and third quarters of the year (59.9%). Snake species was undocumented in 76.2% of records. Pre-hospital first aid was either undocumented (59.1%) or inappropriate such as applying tourniquets (29.3%) and herbal remedies (8.9%). The dominant clinical manifestations were pain (60.8%), swelling/blisters (58.7%), and fang marks (26.9%), while 18.5% had features suggesting envenomation, of whom only 33.3% received antivenom. Overall, only 15.0% of cases received antivenom, while supportive care predominated (intravenous fluids 68.1%, analgesics 64.2%, antibiotics 53.1%). Coagulation testing was rare (20-minute whole blood clotting test 17.3%; INR 12.0%). Overall, 93.4% of patients recovered, 3.1% died, 0.9% were referred to other healthcare facilities, and the rest (2.6%) had missing information on the outcomes. Snakebite management in Northern Uganda is largely symptomatic, with critical gaps in species identification, syndromic assessment, coagulation testing, antivenom utilisation, and pre-hospital first aid practices. Strengthening health-worker training, improving documentation and surveillance, and community education on appropriate first aid are urgently needed to reduce preventable morbidity and mortality.

**Author summary:** Snakebite envenoming is one of the most neglected of the tropical diseases, and its burden falls heaviest on poor, rural communities in sub-Saharan Africa. In Uganda, we know that many people are bitten each year, but very little is known about what actually happens to them once they reach a health facility. We reviewed the records of 227 people treated for snakebite at four hospitals in the Gulu and Arua areas of Northern Uganda between 2017 and 2021. We found that most of those bitten were young people, students and farmers, usually bitten on the leg. Before reaching hospital, many had received harmful first aid such as tight tourniquets or herbal remedies. In hospital, staff rarely recorded which snake was involved, seldom used the simple bedside blood-clotting test recommended by the World Health Organization, and gave the life-saving antivenom to only a small minority of patients, relying instead on painkillers, fluids and antibiotics. Most patients recovered, but some died. Our findings show where care can be improved — through training, better record-keeping, a reliable supply of antivenom, and community education on safe first aid — to prevent needless death and disability from snakebite.

## Introduction

Snakebite envenoming is a life-threatening medical emergency caused by the injection of venom through the bite of a venomous snake or, for some species, defensive spitting of venom into the eyes, leading to severe ocular damage [1, 2]. Globally, an estimated 4.5–5.4 million people are bitten by snakes annually, of which 1.8–2.7 million develop clinically significant envenoming, resulting in 81,000–138,000 deaths and three times as many permanent disabilities, including amputations [3–5]. Clinical sequelae can include neurotoxic paralysis with respiratory failure, venom-induced consumptive coagulopathy and haemorrhage, acute kidney injury, and extensive local tissue necrosis that may necessitate limb amputation [3, 6, 7]. Recognising the magnitude of the problem, in 2017, the World Health Organization (WHO) reinstated snakebite envenoming on the priority list of neglected tropical diseases and two years later (2019), set a global target of halving snakebite-related mortality and disability by 2030 [8].

The burden of snakebite is concentrated in low- and middle-income countries (LMICs), where weak health systems, scarce antivenom, geographical inaccessibility of facilities, and economic vulnerability of rural agricultural and pastoral communities compound the problem [9–11]. Sub-Saharan Africa (SSA) reports an estimated 314,000–435,000 envenomings and 7,300–20,000 deaths annually, accounting for more than 30% of global snakebite-related mortality [12]. The health-economic toll is substantial, as a recent analysis estimated that snakebite imposes catastrophic out-of-pocket costs on households and significant productivity losses on national economies, while antivenom availability remains erratic and unaffordable for most affected populations [13] [14].

In Uganda, the true incidence of snakebite is poorly characterised because of weak surveillance and incomplete facility documentation [9, 15]. A multi-facility survey of 140 health units across six regions in Uganda documented 593 snakebite cases over six months, underscoring a substantial but underestimated burden [10]. Population-based estimates suggest an annual incidence of approximately 101 bites per 100,000 people, with the highest rates in Northern Uganda, where tropical climate, agricultural occupations, and a high diversity of medically important snakes converge [11, 16]. The country’s savannah, wetland, and rainforest ecosystems harbour several clinically important snake species, including the black mamba (*Dendroaspis polylepis*), puff adder (*Bitis arietans*), and various cobras (*Naja* spp.), which cause predominantly neurotoxic, cytotoxic, and haemotoxic envenoming syndromes [17].

Effective hospital management of snakebite largely depends on rapid syndromic assessment, judicious use of the 20-minute whole-blood clotting test (20WBCT) and other coagulation indices, timely administration of polyvalent antivenom in indicated cases, and supportive care for systemic complications [8, 15]. However, knowledge and awareness among healthcare workers and the public remain limited in many LMIC settings [7]. Additionally, pre-hospital practices such as tourniquet application, incision and suction, and herbal remedies persist despite evidence of harm [9, 17, 18]. Studies from Uganda and neighbouring countries have documented variable adherence to clinical guidelines, suboptimal antivenom utilisation, and frequent over-reliance on antibiotics and supportive care [10, 15]. Generating context-specific evidence on how snakebite is actually managed in routine practice is therefore essential to inform clinical guidelines, training, antivenom procurement, and community education. This study, therefore, aimed to describe the demographic and clinical characteristics, pre-hospital first aid practices, in-hospital diagnostic evaluation, treatment, and outcomes of snakebite cases presenting to selected health facilities in Northern Uganda. The findings are intended to inform clinical guidelines, capacity-building for healthcare workers, antivenom supply planning, and public-health interventions aimed at reducing snakebite-related morbidity and mortality in the country.

## Methods

### Study design

This was a facility-based, cross-sectional study using retrospective abstraction of routine clinical records to characterise snakebite cases and assess the quality of in-hospital management in Northern Uganda. The study was reported in accordance with the STROBE (Strengthening the Reporting of Observational Studies in Epidemiology) statement for cross-sectional studies [19].

### Study setting

The study was conducted in Gulu and Arua districts, which are recognised as high-burden areas for snakebite envenomation in Northern Uganda [11, 16]. The region is predominantly rural, with communities largely engaged in subsistence agriculture, livestock keeping, and fishing, which leads to frequent human–snake interactions. Recent population data from the Uganda Bureau of Statistics indicate substantial populations in both districts and their urban centres, including approximately 384,656 residents in Arua City and 233,271 in Gulu City [20]. Population growth and settlement expansion into previously undisturbed ecosystems are likely to increase the frequency of human–snake interactions and demand for snakebite care. Gulu and Arua districts in Northern Uganda experience a substantial snakebite burden and require a health system with adequate capacity to prevent, diagnose, and manage snakebite envenomation [7]. Gulu District, with an estimated population of 443,733, has 65 health facilities comprising 33 government, 15 private, and 17 private-not-for-profit (PNFP) facilities, while Arua District, with an estimated population of approximately 750,000, has 58 health facilities, including 36 government, 17 private, and 5 PNFP facilities [7]. Both districts are served by Regional Referral Hospitals and several general hospitals, Health Centre IVs, Health Centre IIIs, and Health Centre IIs that provide varying levels of emergency and inpatient care.

Epidemiological estimates indicate that Gulu and Arua reported annual snakebite incidences of approximately 104 and 73 cases per 100,000 population, respectively, placing them among Uganda’s high-burden districts [7]. The predominantly rural populations, whose livelihoods depend on crop farming and fishing, coupled with favourable ecological conditions for venomous snakes, contribute to this elevated burden [7]. Despite the existing health infrastructure, snakebite management remains constrained by inadequate antivenom availability, limited training for healthcare workers, insufficient clinical guidelines, weak referral systems, and shortages of essential medicines and laboratory support, underscoring the need to strengthen health system preparedness and capacity for effective snakebite management in these districts [9].

### Site selection and sampling

Gulu and Arua districts serve as significant health service hubs in Northern Uganda, each housing a Regional Referral Hospital, the highest referral facility in its sub-region. These districts are supported by a network of Health Centre IVs, Health Centre IIIs, Health Centre IIs, and private and non-profit facilities that provide preventive, curative, emergency, and referral services to both local populations and neighbouring districts. These facilities play a vital role in managing medical emergencies, including snakebite envenoming. Given that Northern Uganda has the highest incidence of snakebites in the country, Gulu and Arua are particularly suitable locations for evaluating health system readiness and healthcare workers’ capacity in snakebite management.

Four health facilities were purposively selected for this study. We selected these facilities based on the assumption that they possess adequate resources and supplies to effectively handle snakebites, and that they are likely to receive most snakebite cases in the area [21].

In Gulu District, the selected facilities were Gulu Regional Referral Hospital and St. Mary’s Hospital Lacor. In Arua District, the selected facilities were Arua Regional Referral Hospital and Rhema Hospital. Gulu Regional Referral Hospital and Arua Regional Referral Hospital are public Regional Referral Hospitals under the Ministry of Health that provide specialised and referral healthcare services to large catchment populations across the Acholi and West Nile sub-regions, respectively, while also serving as teaching and internship centres [22, 23]. St. Mary’s Hospital Lacor is a large private-not-for-profit (PNFP) hospital that complements public health services by providing comprehensive secondary and tertiary care to patients from Gulu District and neighbouring districts [24]. Rhema Hospital is a private hospital that provides general medical, surgical, obstetric, and emergency services, contributing to healthcare delivery in Arua City and the wider West Nile region [25].

All records with a diagnosis of snakebite captured in the routine registers and clinical files during the period 1 January 2017 to 31 December 2021 were eligible for inclusion. This five-year period was purposively selected to provide an adequate sample of snakebite cases for analysis, capture potential year-to-year and seasonal variations in snakebite occurrence and management, and ensure the use of complete and consistently archived hospital records available at the time of data collection. 2017 as the start year also coincided with WHO recognition of snakebites as a neglected tropical disease (NTD). There were no exclusions based on age or sex.

### Data sources and data collection procedures

Data were abstracted from outpatient and inpatient registers and, where available, individual patient files and clinical notes. Data abstraction was conducted at multiple service points, including adult and paediatric inpatient wards and, where applicable, Intensive Care Units (ICUs), to ensure comprehensive capture of cases across the spectrum of clinical severity.

A semi-structured data abstraction tool was developed from relevant literature [10, 16] and adapted to the Ugandan context. The tool captured (i) sociodemographic characteristics (age, sex, occupation, district, facility ownership); (ii) circumstances of the bite (year, quarter, anatomical site, snake type syndromes/manifestations where documented, time from bite to hospitalisation, where appropriate); (iii) pre-hospital first aid practices; (iv) diagnostic investigations performed, including the 20-minute whole-blood clotting test, INR/PT, complete blood count, renal function, and electrolytes; (vi) in-hospital treatment, including antivenom, antibiotics, corticosteroids, intravenous fluids, analgesics, tetanus toxoid, and supportive measures; and (vii) treatment outcomes (recovery, referral, death). The tool was pre-tested at 1 Public hospital: Kawempe Hospital with capacity to handle snakebites, to ensure clarity and consistency.

Data collection was conducted by trained research assistants holding at least a bachelor’s degree in nursing, clinical medicine, laboratory science, or environmental health, and with prior experience in health research. They underwent a three-day training covering study objectives, data abstraction procedures, ethical considerations, and confidentiality. A reference guide of common clinical terms relevant to snakebite envenomation was provided to minimise misclassification and ensure uniform interpretation. Ongoing supervision and periodic data quality checks were conducted to ensure completeness and accuracy.

### Data management and analysis

Quantitative data were collected electronically using the KoboCollect mobile application (KoBoToolbox, Cambridge, MA, USA), with built-in skip patterns, range checks, and mandatory-field logic to enforce data quality at the point of entry. Data were exported in comma-separated values (CSV) format, cleaned, and imported into Stata version 14 (StataCorp, College Station, TX, USA) for analysis. Descriptive statistics were computed, with frequencies and proportions for categorical variables and means with standard deviations or medians with interquartile ranges (IQR) for continuous variables, as appropriate to the distribution. Where denominators differed because of missing data, the analytic denominator (n) is indicated in the corresponding table. Quality of snakebite data captured remains challenge in Uganda. During data management, records were screened for missing values and data entry errors. Where feasible, missing information was verified against the original data collection forms. Variables with minimal missing data were analysed using complete-case analysis. Findings are presented overall and, where relevant, stratified by district.

### Ethical considerations

Ethical approval for the study was obtained from the Makerere University School of Public Health Higher Degrees, Research and Ethics Committee (MakSPH-HDREC, Ref No: SPH-2021-210) and the study was registered with the Uganda National Council for Science and Technology (UNCST, Ref No: HS2121ES). Administrative clearance was obtained from the District Health Officers in Gulu and Arua districts and from the in-charges of the participating facilities. Since the study was a retrospective review of de-identified routine records, a waiver of individual informed consent was granted by the ethics committee. Patient names and other direct identifiers were not extracted, and each record was assigned a unique study identifier. All electronic data were stored on password-protected, encrypted drives accessible only to the principal investigator. The study was conducted in accordance with the Declaration of Helsinki and the Council for International Organisations of Medical Sciences (CIOMS) International Ethical Guidelines for Health-related Research Involving Humans [26].

## Results

### Characteristics of snakebite cases

A total of 227 snakebite case records were reviewed across the four study facilities. Females accounted for 58.6% (n = 133) of cases, and the median age was 21.0 years (IQR 13–38). Although occupation was not captured in approximately three-quarters of records, where documented, students/pupils (31.3%) and agricultural workers (22.9%) were most frequent. The highest annual proportion of bites occurred in 2019 (30.4%), and the third quarter of the year accounted for the largest proportion of bites (30.4%), followed by the second quarter (29.5%). The lower limb was the most common anatomical bite site (85.9%), and the type of snake was undocumented or unknown in 76.2% of cases (Table 1).

**Table 1.** Demographic, occupational, temporal, and bite-related characteristics of snakebite cases, Northern Uganda (n = 227).

| Variable | Total<br>(N = 227) | District |  |
| --- | --- | --- | --- |
|  |  | Arua (n = 38) | Gulu (n = 189) |
| **Facility ownership** |  |  |  |
| Public | 49 (21.6) | 36 (94.7) | 13 (6.9) |
| Private | 178 (78.4) | 2 (5.3) | 176 (93.1) |
| **Sex** |  |  |  |
|  |  | Arua (n = 38) | Gulu (n = 189) |
| Female | 133 (58.6) | 24 (63.2) | 109 (57.7) |
| Male | 94 (41.4) | 14 (36.8) | 80 (42.3) |
| Age, years; median (IQR) | 21 (13–38) | 17.5 (12–29) | 21 (13–38) |
| **Occupation (n = 192)** |  |  |  |
| Business | 6 (3.1) | 0 (0.0) | 6 (3.8) |
| Herders | 6 (3.1) | 0 (0.0) | 6 (3.8) |
| Agricultural workers | 44 (22.9) | 6 (17.7) | 38 (24.0) |
| Student/pupil | 51 (26.6) | 2(5.3) | 49 (31.0) |
| None mentioned | 77 (40.1) | 25 (73.5) | 52 (32.9) |
| Others (hunter, cleaner, housewife) | 8 (4.2) | 1 (2.9) | 7 (4.4) |
| Missing | 35 | 4 | 31 |
| **Year of bite** |  |  |  |
| 2017 | 47 (20.7) | 2 (5.3) | 45 (23.8) |
| 2018 | 54 (23.8) | 4 (10.5) | 50 (26.5) |
| 2019 | 69 (30.4) | 20 (52.6) | 49 (25.9) |
| 2020 | 12 (5.3) | 6 (15.8) | 6 (3.2) |
| 2021 | 39 (19.8) | 6 (15.8) | 39 (20.6) |
| **Quarter of year the bites happened** |  |  |  |
| First (Jan to March) | 45 (19.8) | 3 (7.8) | 42 (22.2) |
| Second (April to June) | 67 (29.5) | 10 (26.3) | 57 (30.2) |
| Third (July to September) | 69 (30.4) | 10 (26.3) | 59 (31.2) |
| Fourth (October to December) | 46 (20.3) | 15 (39.5) | 31 (16.4) |
| Time in hours from bite to hospitalisation; median (IQR) | 2 (1–6) | 5 (1–9) | 2 (1–6) |
| **Anatomical bite site** |  |  |  |
| Lower limb | 195 (85.9) | 34 (89.5) | 161 (85.2) |
| Upper limb | 27 (11.9) | 4 (10.5) | 23 (12.2) |
|  |  | Arua (n = 38) | Gulu (n = 189) |
| Other body parts | 10 (4.4) | 0 (0.0) | 10 (5.3) |
| **Snake type/syndrome documented (N = 227)** |  |  |  |
| None mentioned / snake not known | 173 (76.2) | 35 (92.1) | 138 (73.0) |
| Cobra or mamba | 9 (4.0) | — | — |
| Puff adder | 15 (6.6) | — | — |
| Vine snake | 2 (0.9) | — | — |
| Green tree snake | 4 (1.8) | — | — |
| Non-venomous snake | 7 (3.1) | — | — |
| Other snake type | 17 (7.5) | — | — |

*IQR, interquartile range, Values are n (%) unless otherwise indicated.*

### Hospital and pre-hospital first aid practices

Documentation of pre-hospital first aid was poor as in 59.1% of cases (n = 133/225) no first aid was recorded. Where first aid was documented, tourniquet application (29.3%) and herbal remedies (8.9%) were the most common practices, while a smaller number of cases involved use of a “black stone” (4.0%), placement of a coin over the bite (2.2%), or other practices including application of raw egg, oral paracetamol, and incising the bite site (5.8%). Circulation, airway, and breathing assessment on arrival was documented in 92.1% of cases (Table 2).

**Table 2.**
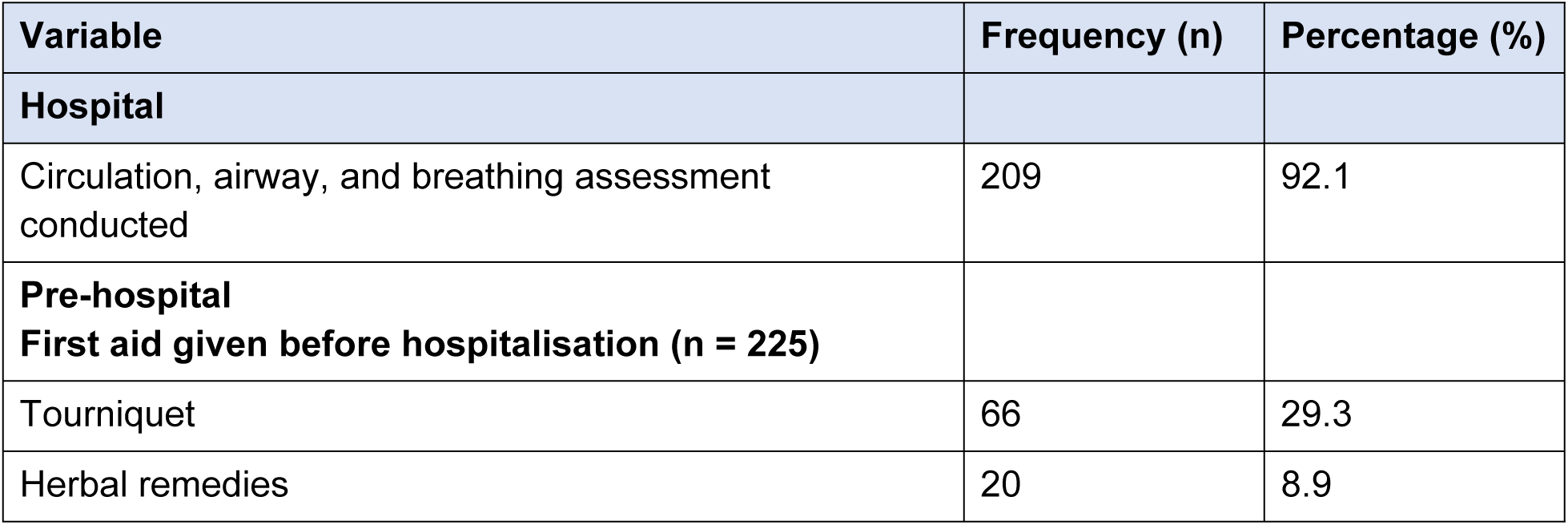
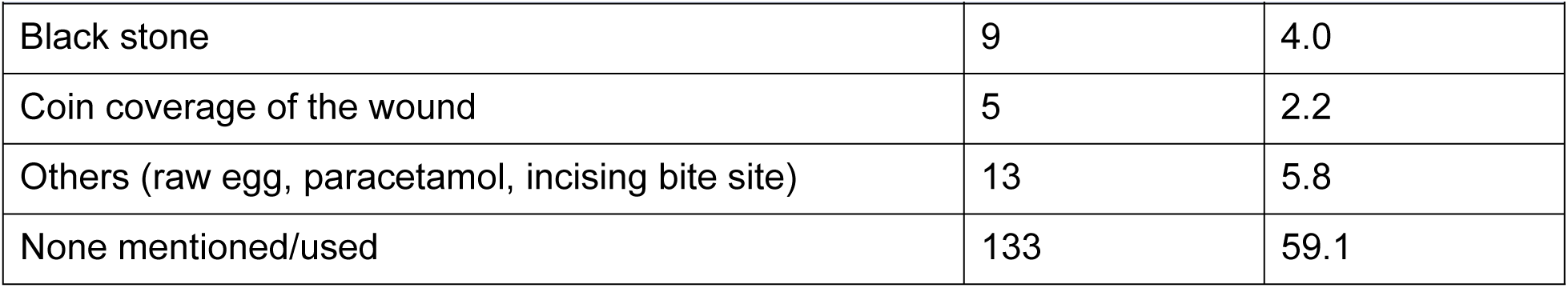
Pre-hospital first aid practices and initial circulation–airway–breathing (ABC) assessment at facilities.

### Clinical manifestations

The most frequently documented clinical features on presentation were pain (60.8%), swelling and/or blisters at the bite site (58.7%), and fang marks (26.9%). Several patients had signs and symptoms suggestive of envenomation, including moderate-to-severe bleeding (7.1%), paralysis of skeletal or respiratory muscles (4.0%), difficulties in breathing (10.6%), excessive salivation or sweating (7.5%), and difficulties in swallowing or speaking. Of these 42 patients, only 14 (33.3%) received snake antivenom (Table 3).

**Table 3.** Clinical manifestations on presentation among snakebite cases (n = 227).

| Sign or symptom on presentation | Frequency (n) | Percentage (%) |
| --- | --- | --- |
| Fang marks | 61 | 26.9 |
| Swelling/blisters at bite site | 156 | 58.7 |
| Pain | 138 | 60.8 |
| Cold extremities | 12 | 5.3 |
| Raised blood pressure | 21 | 9.3 |
| Fever | 22 | 9.7 |
| Laceration | 9 | 4.0 |
| Lymphadenopathy | 1 | 0.4 |
| Mild bleeding | 29 | 12.8 |
| Moderate-to-severe bleeding | 16 | 7.1 |
| Paralysis of skeletal or respiratory muscles | 9 | 4.0 |
| Discolouration of the bite site/limb | 6 | 2.6 |
| Excessive tissue damage | 1 | 0.4 |
| Excessive salivation or sweating | 17 | 7.5 |
| Difficulties in swallowing | 9 | 4.0 |
| Blurred vision | 6 | 2.6 |
| Slurred speech | 3 | 1.3 |
| Difficulty in breathing | 24 | 10.6 |
| Signs of shock | 4 | 1.8 |
| Other features | 52 | 22.9 |
| No documented signs/symptoms | 2 | 0.9 |

### Diagnostic evaluation, treatment, and outcomes

Almost all cases (99.1%) were admitted to the health facility for further management, and 92.5% were admitted for at least 24 hours. Basic vital sign monitoring was relatively common as blood pressure was recorded in 70.4% of cases, pulse rate in 67.7%, and respiratory rate in 54.9%. Whereas laboratory investigations were uncommon. A complete blood count was performed in only 50.9% of cases, snakebite-specific coagulation investigations were markedly underused, with the 20-minute whole-blood clotting test (20WBCT) performed in only 17.3% of cases and INR/prothrombin time in 12.0%. Renal function tests (urea 5.3%, creatinine 5.3%), electrolytes (4.9%), and serum creatine phosphokinase (2.7%) were also infrequent.

Supportive treatment predominated, with intravenous fluids or drugs administered to 68.1% of cases, analgesics to 64.2%, antibiotics to 53.1%, corticosteroids to 45.6%, and tetanus toxoid to 32.3%. Limb elevation was documented in 49.1% of cases, while mechanical ventilation or intubation was documented in 6.2% of the cases. Snake antivenom was administered to only 15.0% of cases overall despite several cases with suggestive envenomation. Of the 227 cases, 212 (93.4%) recovered and were discharged, 7 (3.1%) died, 2 (0.9%) were referred elsewhere for further treatment, and outcome was missing for 6 (2.6%) (Table 4).

**Table 4.** Clinical examination, laboratory tests, treatment and outcomes (n = 227).

| Variable | Frequency (n) | Percentage (%) |
| --- | --- | --- |
| Patient admitted | 224 | 99.1 |
| **Clinical examination** |  |  |
| Pulse rate | 153 | 67.7 |
| Respiratory rate | 124 | 54.9 |
| Blood pressure | 159 | 70.4 |
| Laboratory investigations |  |  |
| INR/prothrombin time | 27 | 12.0 |
| 20-minute whole-blood clotting test | 39 | 17.3 |
| Complete blood count | 115 | 50.9 |
| Blood urea | 12 | 5.3 |
| Creatinine | 12 | 5.3 |
| Electrolytes | 11 | 4.9 |
| Serum creatine phosphokinase | 6 | 2.7 |
| Urinalysis | 6 | 2.7 |
| Electrocardiogram | 4 | 1.8 |
| Blood group testing | 11 | 4.9 |
| None performed | 30 | 13.3 |
| **Treatment administered** |  |  |
| Bed rest | 80 | 35.4 |
| Wound dressing | 35 | 15.5 |
| Reassurance | 3 | 1.3 |
| Sedation | 5 | 2.2 |
| Snake antivenom | 34 | 15.0 |
| Antibiotics | 120 | 53.1 |
| Corticosteroids | 103 | 45.6 |
| Intravenous fluids or drugs | 154 | 68.1 |
| Analgesics | 145 | 64.2 |
| Tetanus toxoid | 73 | 32.3 |
| Mechanical ventilation or intubation | 14 | 6.2 |
| Wound debridement | 1 | 0.4 |
| Limb elevation | 111 | 49.1 |
| Observation of signs and symptoms | 62 | 27.4 |
| Other treatments (antihistamines, oxygen, oral morphine) | 17 | 7.5 |
| **Admitted for $\geq 24$ hours** |  |  |
| No | 6 | 2.6 |
| Not documented | 11 | 4.9 |
| Yes | 210 | 92.5 |
| **Treatment outcomes** |  |  |
| Recovery / discharged | 212 | 93.4 |
| Death | 7 | 3.1 |
| Referred elsewhere for further treatment | 2 | 0.9 |
| No information / missing | 6 | 2.6 |

*INR, international normalised ratio.*

## Discussion

This multi-facility study describes the clinical features, pre-hospital first aid practices, in-hospital management, and outcomes of 227 snakebite cases in two high-burden districts of Northern Uganda. Overall, the findings demonstrate important gaps in the documentation and management of snakebite despite generally favourable patient outcomes. Snakebite predominantly affected children, adolescents, and young adults, with students and agricultural workers representing the most commonly documented occupations, reflecting the vulnerability of economically productive and school-going populations. Documentation of key clinical information, including occupation, snake species, and pre-hospital first aid, was frequently incomplete, limiting comprehensive clinical assessment and surveillance. Furthermore, recommended diagnostic investigations for snakebite envenomation, particularly coagulation and renal function tests, were infrequently performed, and snake antivenom was administered to only one-third of patients with documented envenomation. Although most patients recovered and were discharged, the occurrence of deaths highlights persistent deficiencies in the timely recognition and management of severe envenomation. Collectively, these findings highlight important opportunities to strengthen clinical documentation, improve adherence to evidence-based snakebite management guidelines, ensure timely access to essential diagnostics and antivenom, and enhance the quality of emergency care in high-burden settings, thereby contributing to reductions in snakebite-related morbidity and mortality.

Demographically, most cases were female (58.6%), young, and either students or agricultural workers, mirroring patterns reported in other studies from Uganda and the wider SSA region, where rural occupations, school-age outdoor activity, and household tasks at dawn and dusk drive exposure [27, 28]. The lower limb was the predominant bite site (85.9%), and most bites occurred during the second and third quarters of the year, consistent with peak rainy-season activity of medically important African snakes and increased agricultural labour during planting and weeding seasons [15]. These findings highlight the need for targeted community-based snakebite prevention strategies focusing on high-risk populations, including school-going children and farming communities. They also support the implementation of seasonal risk communication campaigns, promotion of protective footwear and safe farming practices, and strategic prepositioning of antivenom, essential medicines, and trained healthcare personnel in anticipation of periods of increased snakebite incidence.

The predominance of pain (60.8%), local swelling/blisters (58.7%), and fang marks (26.9%), alongside a smaller proportion of systemic features, suggests that most cases presented with mild-to-moderate severity or with dry bites in which little or no venom was injected. A relatively short median delay from bite to facility (2 hours; IQR 1–6) likely limited progression to severe systemic disease and contributed to the high recovery rate. Nevertheless, 18.5% of patients exhibited features consistent with systemic envenomation, including paralysis (4.0%), moderate-to-severe bleeding (7.1%), and respiratory difficulty (10.6%), all of which are clinically important presentations that warrant timely antivenom administration and intensive supportive care [3, 8, 29]. These findings emphasise the need to enhance community awareness in promoting early health-seeking behaviour. It is also crucial to ensure that frontline health facilities are equipped with trained personnel, antivenom, and essential emergency care resources. This will enable them to quickly identify and manage severe envenomation before life-threatening complications arise.

A striking finding was that snake species, or symptoms was undocumented in 76.2% of records. This likely reflects a combination of patients not seeing or being able to describe the snake, limited community familiarity with locally important species, and incomplete clinical history-taking and documentation by healthcare workers [15]. Without syndromic classification suggesting either neurotoxic, cytotoxic, or haematotoxic, distinguishing dry bites and bites from non-venomous snakes from genuine envenoming is difficult, and antivenom decisions may be delayed or inappropriate. Because the offending snake is rarely identified and clinical features alone are seldom diagnostic, WHO recommends a syndromic approach — classifying cases by the pattern of local swelling, the presence of incoagulable blood on the 20-minute whole-blood clotting test, and signs of neurotoxicity — to guide rational, algorithm-based use of polyspecific antivenom. Routine syndromic documentation would therefore both improve individual case management and strengthen surveillance. [8]. Poor documentation also compromises surveillance and antivenom procurement planning, and could affect post-marketing monitoring of antivenom safety and effectiveness [30].

Pre-hospital practices identified in this study echo broader concerns about community knowledge and health-system reach. The frequent use of inappropriate first aid options such as tourniquets (29.3%) and herbal remedies (8.9%), together with the absence of any documented first aid in most cases (59.1%), suggests limited public awareness of evidence-based snakebite first aid (immobilisation of the affected limb, rapid transport to a health facility, and avoidance of tourniquets, incision, suction, and traditional remedies) [31]. Tourniquet application in particular has been repeatedly associated with worsening local tissue necrosis, ischaemic injury, and post-release rebound envenomation WHO guidance is unequivocal that a tight (arterial) tourniquet should never be applied; recommended first aid instead consists of reassurance, immobilisation of the whole patient and especially the bitten limb (for example with a splint or sling), and rapid, passive transport to a facility able to give antivenom. Importantly, when a patient reaches hospital with a tourniquet or other constricting band still in place, antivenom, if indicated, should be started before the band is loosened, because releasing venom sequestered in the occluded limb into the general circulation can precipitate abrupt, life-threatening systemic envenomation. [2, 32]. Reliance on traditional remedies has been linked to delays in formal care and worse outcomes, and frequently reflects geographic and economic barriers to health-facility access as much as cultural preference [9, 33]. These findings support sustained investment in community sensitisation, school health education, and engagement with traditional healers as community gatekeepers.

In-hospital diagnostic evaluation revealed a marked imbalance between basic vital sign monitoring (blood pressure 70.4%; pulse 67.7%) and snakebite-specific coagulation assessment (20WBCT 17.3%; INR 12.0%). Although blood pressure and pulse were documented in most patients, the 20-minute whole-blood clotting test (20WBCT) and INR/prothrombin time were performed in only 17.3% and 12.0% of cases, respectively. This finding is concerning because the 20WBCT is recommended by the World Health Organization and widely adopted across sub-Saharan Africa as a simple, inexpensive bedside test for detecting venom-induced consumptive coagulopathy and guiding decisions on antivenom administration in resource-limited settings [34, 35]. Similar underutilisation of snakebite-specific diagnostic tests has been reported in other African settings, where inadequate training, inconsistent adherence to clinical guidelines, shortages of supplies and laboratory reagents, and limited diagnostic capacity constrain evidence-based management of snakebite envenoming [35–37]. Failure to perform appropriate coagulation testing may result in delayed recognition of systemic envenomation, inappropriate use or withholding of antivenom, and missed opportunities to monitor response to treatment. From a public health and health systems perspective, these findings highlight the need to strengthen implementation of national and WHO snakebite management guidelines through healthcare worker training, routine availability of simple bedside diagnostic tests such as the 20WBCT, improved laboratory support, and regular clinical audit to promote adherence to recommended standards of care.The 20WBCT is also intended to be repeated to monitor treatment: WHO advises that if the blood remains incoagulable six hours after an initial dose of antivenom, the dose should be repeated, and testing continued every six hours until coagulability is restored.

Antivenom utilisation was a particular concern as only 15.0% of all cases and 33.3% of patients with documented envenomation received antivenom. This is consistent slightly higher than the 6.9% reported in a recent multi-region Ugandan retrospective study [15] and the 12.6% reported among severely ill patients in a rural Ugandan ICU series [11]. Underuse of antivenom in SSA is driven by a combination of high cost, which often exceeds the monthly income of affected households, supply-chain disruptions, expiry of stock at the district level, perceived risk of adverse reactions, and limited prescriber confidence [3, 38]. Strengthening antivenom procurement and distribution, ensuring deployment of WHO-prequalified products with demonstrated efficacy against locally important species, and providing protocolised guidance on indications, dosing, and management of acute reactions are critical In line with WHO guidance, antivenom is indicated in all cases of systemic envenoming and of severe local envenoming, should be given as soon as such signs appear, and has no absolute contraindication in life-threatening envenoming; children require the same dose as adults. Intradermal or conjunctival hypersensitivity testing is not predictive of reactions and is no longer recommended, so it should not delay treatment. The potential impact of closing this gap is substantial: early antivenom has historically reduced case fatality from carpet-viper (Echis) envenoming from around 20% to under 3%, underscoring that improving timely, indicated antivenom use is among the highest-yield interventions available in this setting. [2].

The heavy reliance on supportive and adjunctive treatments such as intravenous fluids (68.1%), analgesics (64.2%), antibiotics (53.1%), and corticosteroids (45.6%) indicates that snakebite management in this setting is largely symptomatic rather than definitive. Empirical antibiotic use is widely reported despite evidence that prophylactic antibiotics confer little benefit in the absence of clinical infection Consistent with WHO guidance, antibiotic treatment should be withheld until there are definite signs of infection — such as a hot, reddened, fluctuant local swelling resembling an abscess — or the wound is necrotic; prophylactic antibiotics are appropriate only where the wound has been grossly interfered with or is frankly necrotic. A tetanus toxoid booster, by contrast, is appropriate for all snakebite victims. [34, 39]. Similarly, routine corticosteroid use for envenoming is not supported by current evidence and is recommended only for the prevention or treatment of acute hypersensitivity reactions to antivenom [34]. These prescribing patterns highlight an opportunity for stewardship-focused training, locally adapted clinical algorithms, and structured supervision to align practice with international and regional guidelines [9].

Despite these gaps in clinical management, the overall mortality was 3.1%, comparable to the 2–5% range reported in regional studies and lower than rates reported from facilities with limited critical care capacity [11, 15]. The favourable outcome profile likely reflects predominantly mild envenomations (or none), short delays in presentation, and the protective effect of admission and basic supportive care. Nevertheless, the occurrence of seven deaths and two referrals demonstrates that severe snakebite envenomation continues to result in preventable morbidity and mortality, particularly where timely access to antivenom, advanced supportive care, and critical care services is limited. These findings reinforce the need to strengthen referral systems, ensure the continuous availability of effective antivenom and essential emergency care resources, and build the capacity of frontline health facilities to promptly recognise and manage severe envenomation. Such interventions are critical to reducing preventable snakebite deaths and align with the World Health Organization strategy to reduce snakebite-related mortality and disability through improved health system preparedness and access to quality care.

### Strengths and limitations

This study has several strengths. To our knowledge, it is among the few studies that conducted multi-facility analyses of snakebite management in Uganda, covering both public and private facilities at multiple levels of care, and spanning a five-year period. The use of a structured, pretested abstraction tool, along with trained research assistants and ongoing supervision, strengthens internal validity. The study is reported in accordance with the STROBE statement [19].

Several limitations should be acknowledged. First, retrospective abstraction from routine clinical records is constrained by the quality and completeness of documentation. Indeed, missing data on occupation, snake type, first aid, and outcome were common and may bias estimates. Second, severity classification relied on documented clinical features rather than a standardised severity score, limiting comparison across facilities. Third, only patients who reached a health facility were captured; hence, community deaths, traditional-healer encounters, and bites managed entirely at home were not represented, likely leading to underestimation of true burden and mortality. Fourth, the purposive selection of facilities in two districts limits generalisability to other regions of Uganda. Finally, antivenom batch information, time-to-antivenom intervals, and adverse reaction data were not consistently available.

### Implications for practice and policy

Findings from this study point to several actionable priorities. At facility level, structured in-service training on syndromic recognition, the 20WBCT, antivenom indications and dosing, and management of antivenom reactions is needed. Securing affordable, quality-assured antivenom supply through national procurement and the WHO antivenom prequalification pathway and strengthening referral pathways from health centres to regional referral hospitals are essential. At community level, scaling community-based education on appropriate first aid, snake avoidance, and prompt health-seeking including engagement of traditional healers is critical to reducing pre-hospital delays and harm.

### Conclusion

Snakebite envenomation in Northern Uganda affects predominantly young, female, and agricultural or school-going populations, with most bites involving the lower limb. Care is largely symptomatic, with poor documentation of snake species, limited use of snakebite-specific investigations such as the 20-minute whole-blood clotting test, low antivenom utilisation, and high reliance on adjunctive treatments of questionable benefit. Although overall recovery rates were high, the patterns observed reflect persistent gaps in health-worker capacity, antivenom availability, diagnostic infrastructure, and community first-aid knowledge that are characteristic of snakebite care across sub-Saharan Africa. Targeted health-worker training, secure antivenom supply, strengthened documentation and surveillance, and community education are urgently needed to reduce preventable morbidity and mortality and to advance the WHO 2030 goal of halving snakebite-related death and disability.

## Declarations

### Ethics approval and consent to participate

Ethical approval was granted by the Makerere University School of Public Health Higher Degrees, Research and Ethics Committee (MakSPH-HDREC) [SPH-2021-210], and the study was registered with the Uganda National Council for Science and Technology (UNCST) [HS212ES]. As the study involved retrospective abstraction of de-identified routine clinical records, a waiver of individual informed consent was granted by the ethics committee. Administrative clearance was obtained from the District Health Officers in Gulu and Arua districts and from the in-charges of all participating facilities. The study was conducted in accordance with the Declaration of Helsinki and the CIOMS International Ethical Guidelines for Health-related Research Involving Humans [23].

### Consent for publication

Not applicable.

### Data availability

The de-identified dataset analysed during this study is available from the corresponding author upon reasonable request.

### Author contributions

STW and JBD conceptualised the study, designed the protocol, supervised data collection and analysis, and drafted the manuscript. DM contributed to the study design, supervised field activities, and reviewed the manuscript. STW, AT and SM contributed to data management, and statistical analysis. STW, RKM, and JBI contributed to interpretation of results and critical revision of the manuscript. All authors read and approved the final version of the manuscript and agree to be accountable for all aspects of the work.

## Acknowledgements

The authors are grateful to the District Health Officers and in-charges of Gulu Regional Referral Hospital, St. Mary’s Hospital Lacor, Arua Regional Referral Hospital and Rhema Hospital in Arua for granting access to records. We thank the research assistants (Herbert Simbauni, Pamela Auma, Janet Sherani, and James N Baguma) for their dedication during data collection and the records officers at each facility for their support.

## Use of artificial intelligence tools

No generative artificial intelligence tools were used in the conception, conduct, analysis, or drafting of this manuscript.

